# Adaptation of the PREDIMED Intervention for Medically Underserved Prostate Cancer Patients Through Intervention Mapping

**DOI:** 10.1101/2025.05.15.25327609

**Authors:** Rebekka Garcia, Carrie Daniel, Cindy Hwang, Erma Levy, Matthew Smith, Dalnim Cho, Daniel Frigo, Andrew W Hahn, Hilary Ma, Margaux Robert, Curtis Pettaway, Brian F Chapin, Denise LaRue, Steven Canfield, Jennifer Wargo, Peng Wei, Lorna McNeill, Justin R Gregg

## Abstract

Prostate cancer is the most common non-cutaneous malignancy for men and disproportionately affects those with low socioeconomic status, particularly men from racial and ethnic minority populations. This study describes the development of a culturally tailored Mediterranean diet intervention for medically underserved Black and Hispanic men with prostate cancer, using the Intervention Mapping Adaptation (IM ADAPT) framework. Conducted at a county safety-net hospital in Houston, Texas, which serves a population with high medical needs and low socioeconomic status, the project aimed to ensure the intervention was culturally relevant and evidence-based. A collaborative process was used, involving community scientists and patient stakeholders to identify dietary barriers and preferences, while existing interventions were reviewed for cultural fit. Guided by the six steps of the IM ADAPT framework, stakeholder feedback was incorporated throughout the adaptation process. The result was a culturally adapted intervention that included tailored dietary modifications, food provision strategies, and educational materials specifically designed for Black and Hispanic men. The finalized intervention is ready for pilot testing and may serve as a model for adapting evidence-based interventions to address health disparities in other racial and ethnic minority populations affected by cancer.

## Introduction

Prostate cancer (PCa) is the most common non-cutaneous malignancy for men. PCa diagnosis and subsequent treatment can lead to significant risks of morbidity and mortality, including changes in urinary and erectile function (Hamdy et al., 2023; Resnick et al., 2013) as well as other quality of life measures (Huang et al., 2019). It is established that low socioeconomic status (defined as living in an area with greater than or equal to 20% of the population below the poverty line) increases PCa mortality in men of all races and ethnicities, most notably in Black men (Ward et al., 2004). Furthermore, uninsured men with PCa have a mortality rate almost twice that of those with insurance (Niu et al., 2013). Despite these disparities, minority PCa patients have been underrepresented in PCa clinical trials (Balakrishnan et al., 2019).

Dietary interventions are an attractive means to modify risk of PCa progression given their noninvasive nature, impact on cardiovascular risk factors (many men with localized tumors will die of cardiovascular disease rather than PCa (Bill-Axelson et al., 2018)), and potential modification of markers of PCa progression. However, the few existing interventions that have been developed for men with PCa have not yet demonstrated cancer-specific benefits in any sociodemographic group. Further, none have been specifically designed for medically underserved populations, and few studies have evaluated barriers to dietary change.

Our group has shown that adherence to a Mediterranean dietary pattern, which is rich in fruits, vegetables, whole grains, and olive oil and has established cardioprotective effects (as shown in the landmark PREDIMED trial (Estruch et al., 2006)), may be associated with decreased risk of PCa progression in men whose disease is actively monitored (Gregg et al., 2021). A Mediterranean diet may have beneficial effects on the gut microbiome (Martínez-González et al., 2015a), inflammation (Chrysohoou et al., 2004; Estruch et al., 2006; Guasch-Ferré et al., 2017), lipid levels (Doménech et al., 2014a; Martínez-González et al., 2015a), and metabolic health (Doménech et al., 2014a; Martínez-González et al., 2015a), all of which are increasingly recognized to impact outcomes across the cancer continuum. Our group has also shown that levels of certain lipid metabolites are associated with PCa progression and are potentially responsive to a strict, short-term Mediterranean diet-based intervention (data unpublished). Therefore, Mediterranean diet-focused interventions may impact rates of disease progression while mitigating risks of cardiovascular disease and other comorbidities. However, Mediterranean diet intervention has not been adapted for medically underserved groups.

There are many established barriers to dietary change, including among medically underserved populations. These barriers include: 1) lack of knowledge about Mediterranean diet foods (social construct theory), (Hardin-Fanning, 2013) 2) resistance to dietary change (theory of planned behavior; health belief model), (S. E. Moore et al., 2018) 3) concern that dietary components would not be appealing (theory of planned behavior), (Kretowicz et al., 2018) and (Kretowicz et al., 2018) 4) and social determinants of health (theory of planned behavior; health belief model) (S. E. Moore et al., 2018).

Here, we detail the process used to develop and culturally adapt an existing evidence-based dietary intervention for low-income, ethnic and racial minority men with PCa. To that end, we define the decision-making process of the adaptation and describe the strategies used to create the intervention, adapted for behavioral modification in the target population that addresses barriers to dietary change in this group.

## Methods

### Study Design

We aimed to develop a Mediterranean diet intervention for low-income, racial and ethnic minority men with PCa. We began with the creation of a logic model of change, incorporating factors associated with dietary change, PCa progression, and behavioral theory (**Figure 1**). The development of a Mediterranean diet intervention was based on the Intervention Mapping Adaptation (IM ADAPT) framework (Highfield et al., 2015), a systematic approach for evidence-based interventions to better fit the needs of specific populations. It builds on Intervention Mapping (IM) (Fernandez et al., 2019) and incorporates Adaptation (ADAPT) principles (G. Moore et al., 2021) to ensure interventions remain both evidence-based and culturally relevant. IM ADAPT is a six-step method for adaptation which guides program planning in developing an evidence-based intervention (Cho et al., 2020). The steps include: (a) needs assessment by describing the problem and organizational capacity; (b) searching for evidence-based interventions that have been developed for the target population; (c) assess the behavioral and environmental fit of the intervention and plan adaptations for the target population; (d) implement the adaptation of intervention materials and assessment tools (e) plan for the implementations; and (f) plan for the evaluation of the intervention. This framework balances scientific rigor with cultural relevance, making it useful for tailoring dietary interventions for minority populations. We performed all steps of the IM ADAPT framework, resulting in the intervention described herein.

**Figure 1.**
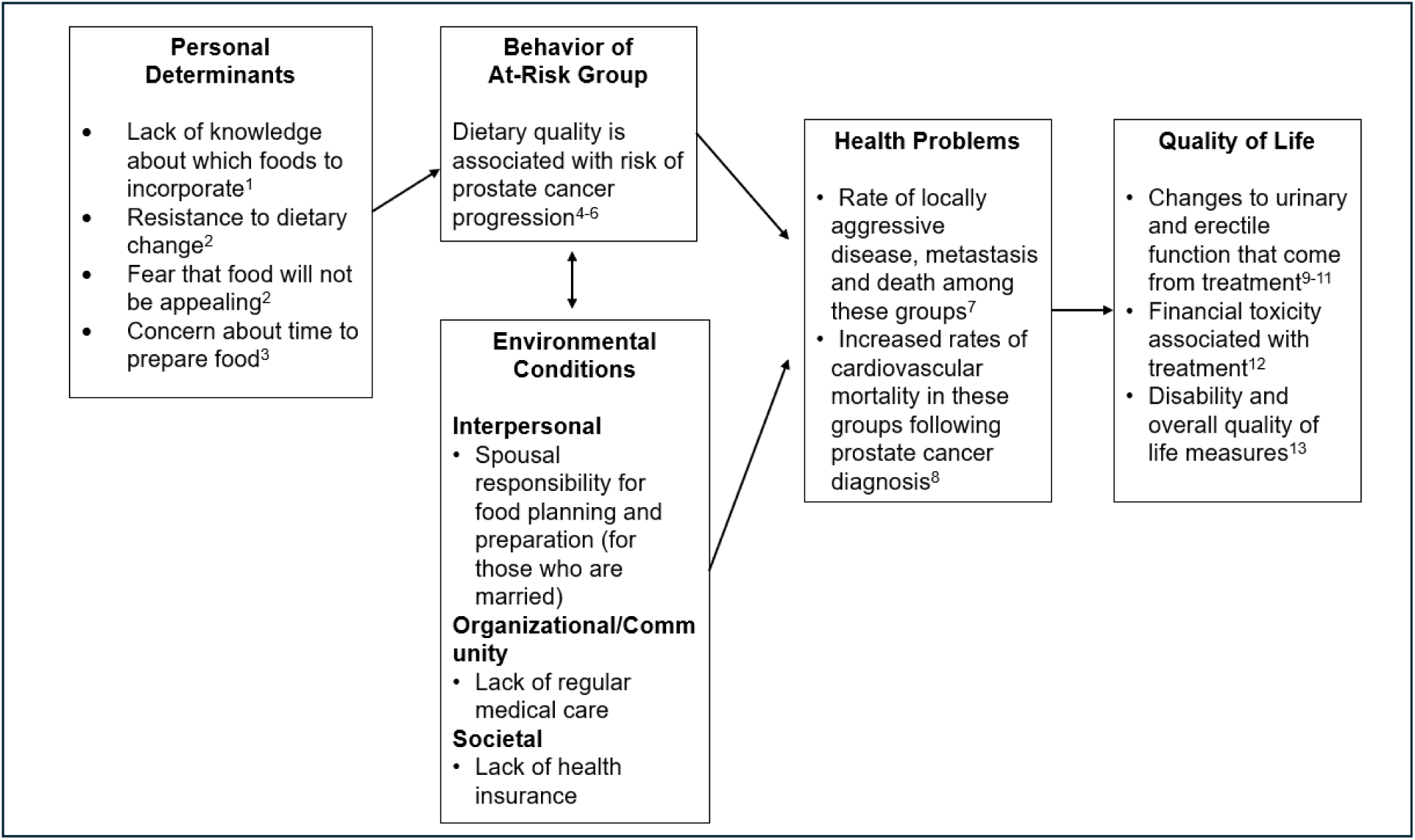
Logic Model of Behavior Change. Logic Model developed to address the risk of development of aggressive prostate cancer and treatment-related changes to quality of life among Black Men and men of other races/ethnicities with low socioeconomic status who are 50 to 70 years old. ^1^Hardin-Fanning, 2013; ^2^ Moore et al., 2018; ^3^ Kretowicz et al., 2018; ^4^Bosire et al., 2013; ^5^Yang et al., 2015; ^6^ Möller et al., 2013; ^7^ Siegel et al., 2021; ^8^Ward et al., 2004; ^9^Hamdy et al., 2016; ^10^Bill-Axelson et al., 2018; ^11^Resnick et al., 2013; ^12^Stone et al., 2021; ^13^Huang et al., 2019

## Results

### Application of the IM ADAPT Framework

#### Step 1: Organizational Capacity and Needs Assessment

The study was developed by a physician scientist-led team at The University of Texas MD Anderson Cancer Center (MDACC) and Harris Health Lyndon B Johnson (LBJ) Hospital in Houston, TX. MDACC is a multi-specialty, National Cancer Institute-designated comprehensive cancer center. LBJ Hospital is a county safety-net facility that serves a large medically underserved population. The multi-disciplinary research team was headed by a physician scientist with expertise in health disparities and urologic oncology and included nutritional epidemiologists, registered dieticians, and statisticians.

A needs assessment was performed by evaluating preliminary data from men undergoing biopsy at LBJ Hospital. We began with data in which we assessed Mediterranean diet patterns among a broader population of men with localized PCa seen at a tertiary cancer center. We included 410 men with localized PCa on active surveillance (**Supplementary Table 1**) who completed the VioScreen questionnaire, and a Mediterranean diet score was calculated using the MEPA I instrument (Cerwinske et al., 2017). The population averaged a score of 4 on a scale of 0-16, indicating low adherence to the Mediterranean Diet pattern. These findings, coupled with our clinical experience and data indicating that low-income PCa survivors seen at a county facility exhibit poor adherence to dietary recommendations (Haymer et al., 2020) led us to conclude that Mediterranean diet adherence is likely low in our target population.

Following this, we met with trained community scientists and presented intervention goals and proposed study framework. Community scientists were asked for their initial reactions, thoughts on dietary interventions, ideas to increase the success of and potential barriers to the intervention. We presented a general framework in which we would complete a study focused on Mediterranean diet change based on an evidence-based intervention that would: 1) leverage existing resources within the Harris Health system 2) utilize an evidence-based patient education program and 3) rely on patient feedback regarding design and enrollment. We also planned to create two separate educational programs, one for Black men and one for Hispanic men, given evidence that Hispanic men may have lower adherence to a Mediterranean dietary pattern than Black or non-Hispanic White men (Harmon et al., 2015). Community scientist feedback was generally positive, with unanimous support to move forward with the intervention. Comments included that the study team needed to consider providing lessons on how to use some food items to create healthier meals, and identification of food substitutions. Because this intervention is aimed at the medically underserved, the community scientists warned that all patients may not be able to afford all the ingredients to create meals. It was suggested that we have a community stakeholder support the study and promote it to create trust in the community. Barriers mentioned included that some people are not familiar with the food items in a Mediterranean diet and are unsure how to cook them. Food preparation instructions and ideas were suggested to meet this barrier. To encourage enrollment in our study, the community scientists suggested sending extra food to share with the family, including recipes and videos, sending pre-prepared items to add to a meal (e.g. nuts), while some suggested providing memberships to meal delivery services or gift cards to “healthy” food stores (though others met this suggestion with skepticism). Finally, the community scientists noted that education on healthy diet can create a generational change by passing on appreciation for healthier food choices and that it was important to include the whole family in the intervention.

#### Step 2: Evidence-Based Intervention Search

The Mediterranean diet is characterized by an intake of legumes, whole grains, vegetables, fruits, fish, nuts, and olive oil and is well known for its anti-inflammatory (Chrysohoou et al., 2004; Estruch et al., 2006; Guasch-Ferré et al., 2017) and lipid-lowering (Doménech et al., 2014b; Martínez-González et al., 2015b) effects. We considered several evidence-based interventions to inform our adaptation. The Med-South Lifestyle Program was developed 30 years ago and has been used in many research studies and community interventions (Samuel-Hodge et al., 2022). It combines Mediterranean and Southern food culture, delivered through monthly counseling sessions, phone-call check-ins, and a program manual and cookbook. Four monthly education models focus on various aspects of health: taking medications, smoking cessation, healthy eating, and physical activity. The PREDIMED diet is a well-established intervention in which 7447 Spanish men and women (55-80 years old with high cardiovascular disease risk) were randomly assigned to diet groups. Patients who were randomized to one of the two Mediterranean diet groups (who received behavior-focused dietitian counseling and diet supplementation with nuts or olive oil) had a lower incidence of cardiovascular events when compared to a control (low fat diet) group (Estruch, 2006). The intervention included baseline and quarterly individual and group educational sessions, provision of nuts or olive oil (to those randomized to Mediterranean diet), and motivational interviewing techniques (Miller, 2023). **Table 1** describes the components of both interventions. After careful consideration of program strengths and our objectives, we selected the PREDIMED intervention due to its focus centered around Mediterranean diet change and strong evidence in preventing cardiovascular disease, which is a leading cause of death among men with PCa (Berglund et al., 2011). While we additionally appreciated the well-rounded approach of the Med-South program, we also noted only a modest dietary change achieved in the pilot, including an average increase of 0.4 servings of nuts and 0.9 servings of fruit and vegetable per day (Samuel-Hodge et al., 2022).

**Table 1.** Comparison of Two Dietary Interventions for Adaptation: The Med-South Lifestyle Program and the PREDIMED Diet.

|  | **Med-South Lifestyle Program** | **PREDIMED diet** |
| --- | --- | --- |
| Target population | Men and women in North Carolina, USA, 18-80 | Men and women in Spain, 55-80 at high cardiovascular risk |
| Intervention domains | Dietary change, physical activity, medication compliance, smoking cessation | Dietary change |
| Dietary change focus | Fruit and vegetable intake | Mediterranean diet with olive oil and walnuts |
| Study Design | Hybrid implementation-effectiveness study of monthly community health worker-delivered behavioral intervention promoting dietary change (based on Mediterranean diet), increased physical activity and other health measures. | Randomized, phase III study with two intervention (Mediterranean diet and olive oil provision and Mediterranean diet and walnut provision) and one control group (low fat diet) that included quarterly dietitian visits with motivational interview techniques. |
| Number of patients | 255 (72.2% female) | 7447 (57% women) |
| Intervention length | 4 months | 4.8 years |
| Years of enrollment | 2016-2019 | 2003-2009 |
| Published outcomes | Number and demographics of participants recruited and retained; blood pressure; compliance; select dietary change measures.<sup>1</sup><br>Dietary change noted increase of 0.8 daily servings of fruits and vegetables and physical activity by 45 minutes/week. | Composite of major cardiovascular events (myocardial infarction, stroke and/or death from cardiovascular causes). Decrease in risk noted among Mediterranean diet group compared to control (HR 0.69 95% CI 0.53-0.91).<sup>2</sup><br>Increase in MEPA I score in Mediterranean diet intervention groups. Decrease in composite of cardiovascular disease risk. |

<sup>1</sup> Samuel-Hodge CD, Ziya Gizlaice, Allgood SD, et al., Am. J. Health Promot. 2022
<sup>2</sup> Estruch R. Ann. Intern. Med. 2006

#### Step 3: Fit Assessment and Adaptation Planning

To assess the fit of the intervention, six focus groups (n=31), three in Spanish (with Hispanic men and their spouses; n=18), and three in English (with Black men and their spouses; n=13) were conducted at LBJ Hospital over five months. Participants were men who had been diagnosed with localized PCa over the previous two years. Spouses and significant others (defined as a spouse, significant other or family member as identified by the patient) were also invited to participate. Spouses and significant others were included due to their established role in food provision and preparation (Flagg et al., 2014). Focus groups were convened to answer central questions about current dietary habits, Mediterranean diet knowledge and barriers to dietary change (in general). The results of this will be published separately, though preliminary thematic analysis suggests themes including the importance of physician recommendations and knowledge, relationships with family and friends, and current food habits.

An additional objective of the focus groups was to gain feedback on proposed educational materials, dietary structure and interventions, all of which were based on the PREDIMED study. Noting that our study population (men and their spouses at LBJ Hospital) was very different from those in PREDIMED (predominantly older individuals from Spain), and the earlier time frame in which (and available funding for) pilot studies following intervention completion, we planned to draft new materials to be given during a weekly time frame (rather than sporadically over months to years). Our study team, led by expert dietitians, drafted educational materials in English and Spanish containing lessons on tenants of the Mediterranean diet (e.g. fruits and vegetables, whole grains), which were prioritized given preliminary feedback that dietary knowledge may be an important barrier to change in this population. In addition, given established concerns that food palatability may act as a barrier to change (Kretowicz et al., 2018), we additionally prepared lists of proposed food substitutions believed to be culturally specific, obtaining specific feedback about each. The final list of substitutions for inclusion in intervention education and materials is listed in **Table 2**.

**Table 2.** Food substitution examples discussed in focus groups.

| **Original food** | **Substitution** |
| --- | --- |
| ½ cup butter | 1 ½ ground flaxseed |
| Mayonnaise or butter | Slice avocado |
| Creamy dressing | Olive oil vinaigrette |
| Flour tortillas | Corn or whole wheat tortillas |
| Full fat dairy | Low- or no-fat dairy |
| Ground beef | Plant protein |
| Fried fish | Baked fish |
| Pan crust and/or meat pizza | Thin crust and/or veggie pizza |

#### Step 4: Adaptations

Using this information, culturally relevant educational materials were then finalized in Spanish and English (**Appendix 1 & 2**). Materials consisted of 8 lessons that instruct and inform participants into the foundations of the Mediterranean diet and how to implement them into their daily lives. The intervention itself, along with educational materials, focuses on addressing established barriers to Mediterranean diet intervention: 1) lack of knowledge about Mediterranean diet foods (social construct theory), (Hardin-Fanning, 2013) 2) resistance to dietary change (theory of planned behavior; health belief model), (S. E. Moore et al., 2018) and 3) concern that dietary components would not be appealing (theory of planned behavior) (Kretowicz et al., 2018). Additionally, the inclusion of motivational interview techniques by study dietitians was considered an important aspect of the PREDIMED intervention to be included in the adapted intervention given it will be used to support the outcome expectations. Finally, spouses and significant others are included in the intervention given their role in food preparation, determined through stakeholder feedback and community scientist feedback and data that social support is a relevant behavioral determinant among minority men with PCa (Raber et al., 2023). In addition, we prioritized food provision given that many patients at LBJ live in neighborhoods considered food deserts (Raber et al., 2023) and patient and community scientist feedback. The intervention will provide provisions in two ways: first, through existing collaboration with the Harris Health system’s innovative food pharmacy, through which fresh fruit, vegetables, and other foods will be provided every other week during the 8-week study. Furthermore, the food pharmacy will provide education about food preparation and resources regarding ongoing food support such as through the SNAP program. Second, olive oil and walnuts will be provided during the 8-week intervention, consistent with the PREDIMED intervention (Estruch, 2006). A quantity will be provided to supplement both the patient and his family’s diet during the 8-week intervention, while personalized education about diet, food choices, and preparation is occurring. During the intervention, dietitians will perform weekly nutritional counseling with motivational interview techniques. Finally, a website developed at MD Anderson <u>At The Table - MD Anderson Cancer Center</u> (https://atthetable.mdanderson.org) that contains recipes (including those specific to Mediterranean cooking) in both English and Spanish will be provided and explained by study dietitians. Participants will be compensated with $25 gift cards for each of 6 visits based on feedback regarding issues such as transportation and time-related costs.

#### Step 5: Implementation Planning

All study activities will take place at LBJ Hospital. Patients who are self-identified Hispanic and/or Black and who have been diagnosed with localized PCa in the previous twelve months or who are currently managed on active surveillance (with their initial biopsy being less than 5 years from enrollment) will be included.

The intervention (**Figure 2**) will be delivered by a physician overseeing care at the LBJ urology clinic. Spanish-speaking, bilingual study coordinators will collect food questionnaire data and meet with patients at biweekly clinic visits. Registered dietitians will provide nutritional counseling weekly by phone, and patients will visit the Food Farmacy which is on-site at LBJ hospital following their biweekly visits to receive additional education and food. Olive oil and nuts will be provided during clinic visits at weeks one and four.

**Figure 2.**
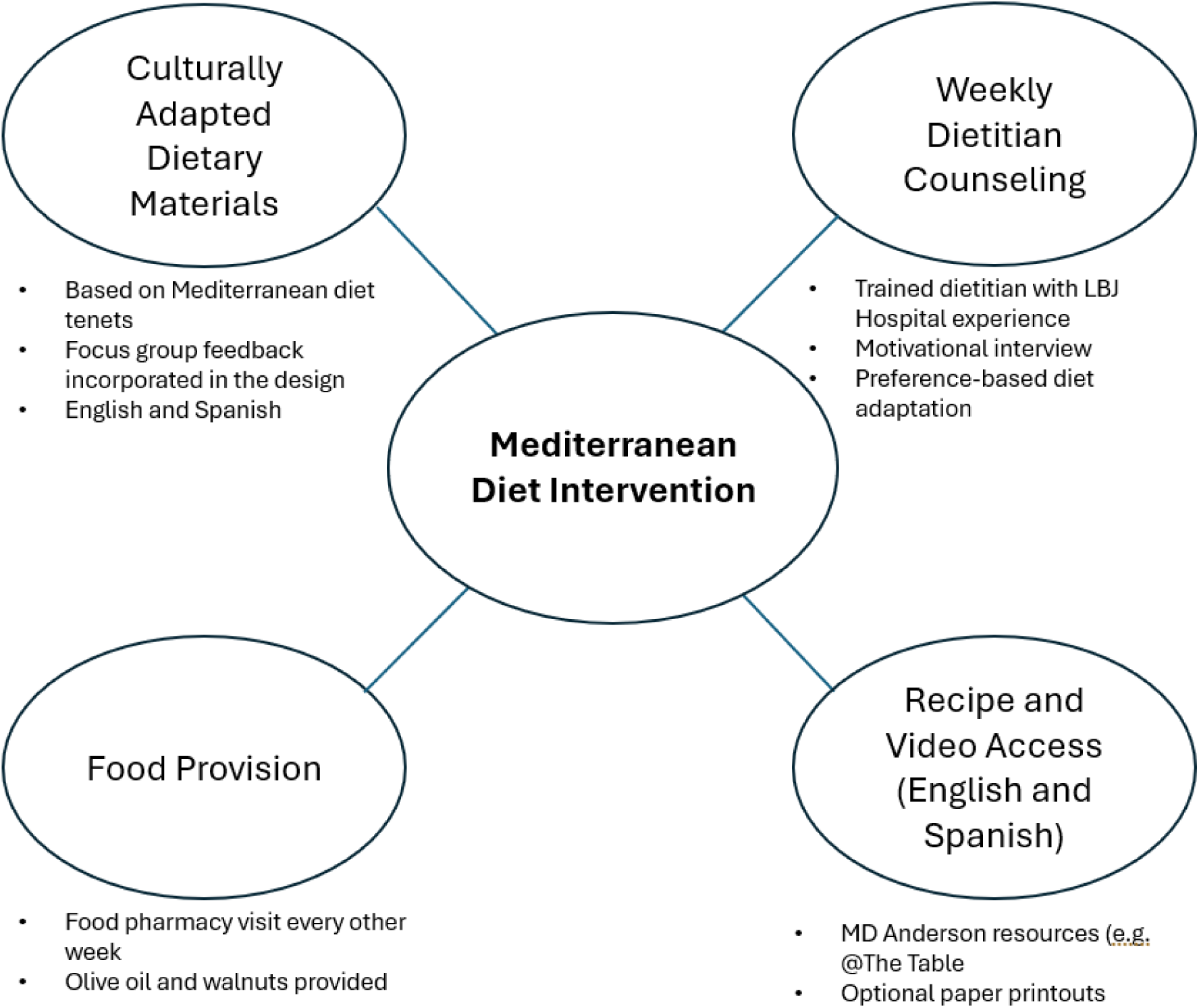
Summary of Mediterranean diet intervention. Each circle is an intervention domain, offering examples of use and patient interaction. Educational materials and weekly dietitian counseling (including motivational interview techniques) will be provided by MD Anderson’s Bionutrition Research Core. Patients will also access LBJ’s food pharmacy, which provides cancer patients with fresh fruits and vegetables at no cost in addition to education about food preparation and the use of resources such as the SNAP program. Nuts and olive oil will be provided, consistent with the PREDIMED intervention.^22^

In terms of plan adopters (Bartholomew Eldredge et al., 2016), Harris Health leadership and the food pharmacy were identified as important groups needed to complete this pilot, and their support has been secured. Program implementers will start on the clinical side, from which participants will be recruited. These include participating physicians as well as advanced practice providers and trainees who aid in clinical care. Finally, program maintenance will be dependent on both program adopters and implementers. It will be key to demonstrate to clinic leadership and staff that the study falls within the timing and space of normal clinical encounters, therefore offering potential benefit to study participants without disrupting clinical workflows.

Adherence to the protocol will be measured with a food frequency questionnaire as well as discussions with their dietitians and the study coordinator.

#### Step 6: Evaluation planning

We propose a one group, quasi-experimental design to determine the feasibility of Mediterranean diet intervention grounded within the ORBIT model (Bartholomew Eldredge et al., 2016) (**Figure 3**). The **primary outcome of the study will be dietary adherence**, defined as the quantitative increase in Mediterranean diet score on the MEPA III questionnaire (Cerwinske et al., 2017) (obtained using data from the VioScreen surveys at baseline and 6 weeks). Secondary outcomes include compliance (percentage of food provided that was consumed, with over 80% considered compliant), enrollment (completion of 25 patient enrollment within 18 months considered successful), retention (completion of the full study duration and at least 80% of counseling sessions), tolerability (based on participant feedback during dietitian counseling sessions), and multiple measures of quality of life listed below. If benchmarks are not met, the bidirectional nature of the ORBIT model (Powell et al., 2021) (**Figure 3**) will enable us to take a step back and address study design or recruitment strategies to optimize feasibility before advancing.

**Figure 3.**
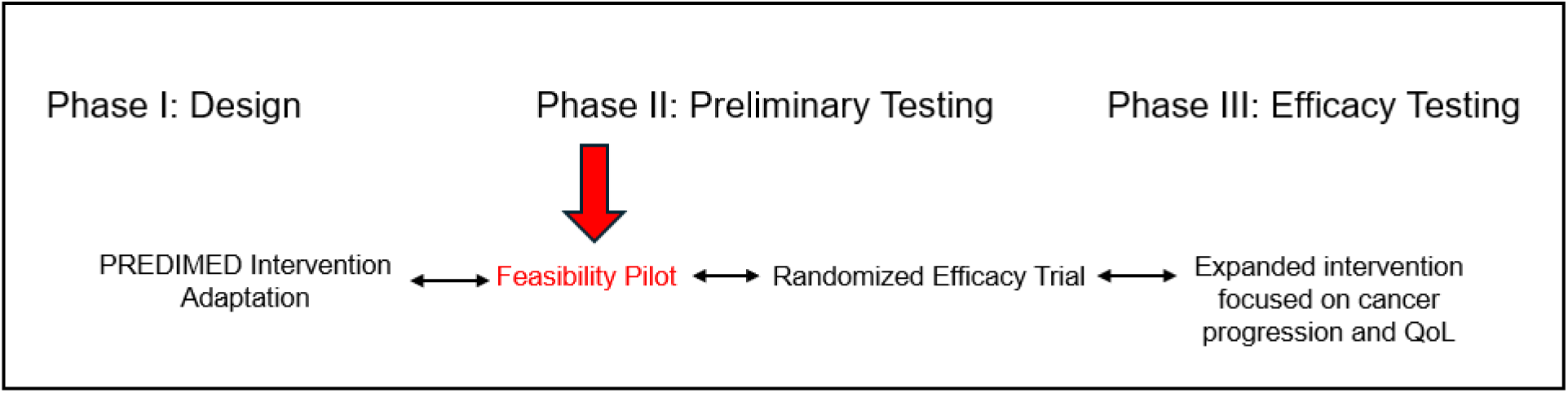
Use of the ORBIT model of behavioral interventions in developing a Mediterranean diet intervention for medically underserved men with Prostate Cancer. Three phases of the ORBIT model are shown. Adaptation of the PREDIMED intervention for medically underserved men with PCa is described in this manuscript. The currently proposed study is a feasibility pilot that will be used to determine feasibility of enrollment, recruitment and retention is highlighted in red. Successful completion of the proposed feasibility pilot will allow progression to a randomized efficacy study of the association between diet change and clinical (e.g. progression) and/or relevant biomarker changes. ORBIT model progression is bidirectional; thus, we can return to the design phase if intervention is not feasible. The long-term objective is to impact PCa outcomes and quality of life (QoL), highlighted in Phase III.

Regarding quality of life measures, we will assess urinary and erectile function (EPIC-26 (Wei et al., 2000)) andQoL (EQ-5D-5L (Herdman et al., 2011)). We will additionally collect biospecimens (urine, blood and stool) for use in additional studies that allow future studies designed to evaluate changes in established markers of PCa progression and cardiovascular disease that may be modified by dietary change. Finally, after dietary intervention we will assess sustainability through a 6-month VioScreen assessment (Kristal et al., 2014).

## Discussion

We describe an evidence-based, culturally specific adaptation of the PREDIMED dietary intervention for medically underserved Hispanic and Black men with PCa (and their spouses), guided by the IM ADAPT framework. Several observations became evident during this adaptation process. First, while the adaptation was based on the PREDIMED study, its tailoring for this specific population for a different disease required a comprehensive process. A multi-disciplinary research team was assembled to ensure the adaptation maintained scientific integrity through a thorough needs assessment, establishment of a logic model, extensive investigation of prior evidence-based interventions, evaluation of the fit of the adaptation, creation of the adaptation, and implementation and evaluation of the new intervention. Community feedback was additionally incorporated at multiple time points.

Notably, the adaptation process was not linear. Lessons learned in the assessment process guided changes in the adaption (such as through modification of food substitutions (**Table 3**) and provision of nuts and olive oil for participants and family members). Further, community feedback obtained following focus group feedback consistently identified food cost as a barrier to Mediterranean diet change, leading our group to prioritize two methods of food provision (direct provision of nuts and olive-oil and biweekly visits to the on-site food pharmacy) and to provide gift cards as compensation for participation. Furthermore, concerns regarding self-efficacy in preparing Mediterranean food led us to increase emphasis on preparation discussions with the dietitian and food pharmacy staff. Despite these identified barriers, all men were receptive to the proposed intervention and had a positive response.

The present study has several limitations. Though we reached saturation in the focus groups, there is a possibility that their experiences are not representative of all medically underserved men with PCa. Furthermore, this intervention is adapted for a specifically low-income, ethnic and racial minority population and may not be generalizable to all men with PCa.

To the authors’ knowledge, this is the first Mediterranean diet intervention explicitly informed by a cultural adaptation process for medically underserved groups of men with PCa in the United States. This adaptation may serve as a foundation for future interventions for men with PCa, with particular relevance to racial and ethnic minority men and those at heightened risk for adverse outcomes. Mediterranean diet change offers the potential to impact both rates of cardiovascular disease (Estruch, 2006) and PCa progression risk (Gregg et al., 2021), offering the promise of improved outcomes and quality of life among at-risk men diagnosed with PCa. Pilot testing and further study of this multi-disciplinary intervention is needed to determine the success of this intervention, after which it may be used as part of future large-scale clinical trials designed to evaluate the impact of Mediterranean diet change on oncologic outcomes and quality of life.

This study describes the systematic application of the IM ADAPT framework to adapt an evidence-based diet intervention for men with PCa, with particular attention to the needs of racial and ethnic minority men who experience disproportionate PCa burden. The adaptation process was guided by theoretical foundations, empirical evidence, and stakeholder input, resulting in a culturally relevant and contextually appropriate intervention. This work highlights the importance of balancing fidelity to evidence-based practices with necessary adaptations to enhance fit, feasibility, and acceptability in underrepresented populations. The intervention adaptation process described here offers a replicable model for researchers and practitioners aiming to address health disparities through culturally responsive intervention design. Future work will focus on evaluating the adapted intervention to assess its effectiveness in improving dietary behaviors, with the plan to complete future studies focused on improving PCa outcomes in diverse populations.

## Data Availability

All data produced in the present work are contained in the manuscript.

## Notes

Conflicts of Interest: JRG – Consultant/Advisory board: Bayer and J&J, AWH – Consultant/Advisory board: Janssen, Intellisphere, AVEO, Exelixis, Eisai, Pfizer, and Tolmar. Honoraria: Medscape, Binaytara Foundation, Projects in Knowledge, Curio Science, Dava Oncology, and Mashup Media. Institutional research funding from Bayer, Eisai, and Halda Therapeutics, DEF – Familial relationship: March Biosciences, Biocity Biopharmaceuticals, and Barricade Therapeutics

### Competing Interest Statement

JRG: Consultant/Advisory board: Bayer and J&J
AWH: Consultant/Advisory board: Janssen, Intellisphere, AVEO, Exelixis, Eisai, Pfizer, and Tolmar. Honoraria: Medscape, Binaytara Foundation, Projects in Knowledge, Curio Science, Dava Oncology, and Mashup Media. Institutional research funding from Bayer, Eisai, and Halda Therapeutics
DEF: Familial relationship: March Biosciences, Biocity Biopharmaceuticals, and Barricade Therapeutics

### Funding Statement

This study did not receive any funding.

### Author Declarations

IRB of University of Texas MD Anderson Cancer Center gave ethical approval for this work.

