## Supplemental Table 1 and Appendices for "Adaptation of the PREDIMED Intervention for Medically Underserved Prostate Cancer Patients Through Intervention Mapping"

**Supplementary Table 1. Baseline characteristics of men with localized prostate cancer on active surveillance by tertiles of the Mediterranean Diet score (MEPA I).**

| Characteristic (N=410 total) | Low: 0-3 (n=141) | Medium: 4-5 (n=171) | High: 6-9 (n=98) | P Value |
| --- | --- | --- | --- | --- |
| Age, mean (SD), y | 62.8 (8.5) | 64.9 (8.0) | 65.8 (8.6) | .02 |
| Race, No. (%) |  |  |  | .61 |
| White | 113 (80.1) | 147 (86.0) | 80 (81.6) |  |
| Black | 14 (9.9) | 12 (7.0) | 7 (7.1) |  |
| Other/unknown | 14 (9.9) | 12 (7.0) | 11 (11.2) |  |

**Appendix 1. Mediterranean Diet Intervention Curriculum (English). These materials were translated to Spanish for Hispanic patients (data not included).**

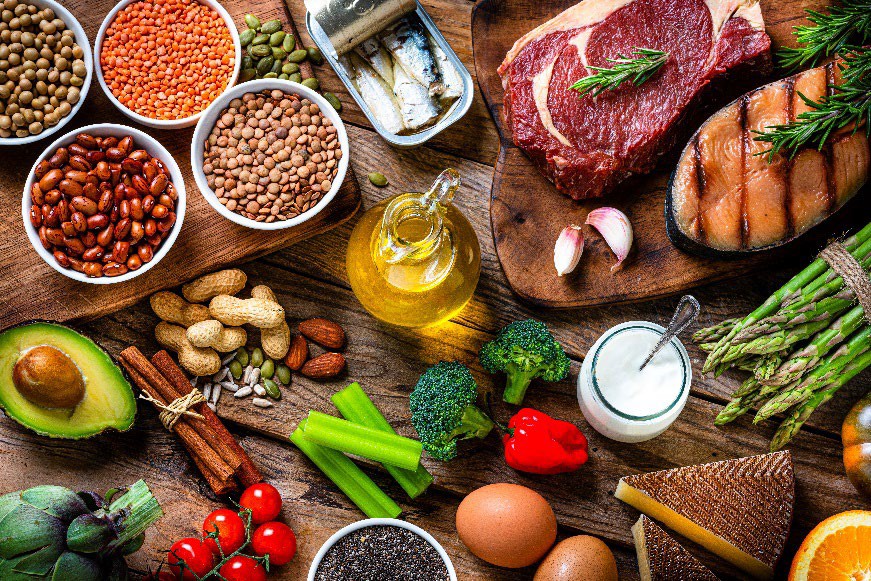

### Lesson 1: The Mediterranean Diet Eating Pattern

The Mediterranean diet is a healthy, balanced diet. It includes a variety of flexible choices and focuses on “whole foods” that are less processed and provide complete nutrition.

#### Health benefits:

- Lowers your risk of cardiovascular diseases
- Helps to achieve a healthy body weight
- Supports healthy blood sugar levels, blood pressure, and cholesterol levels
- Reduces inflammation
- May lower your risk of worsening prostate cancer and risk of other cancers

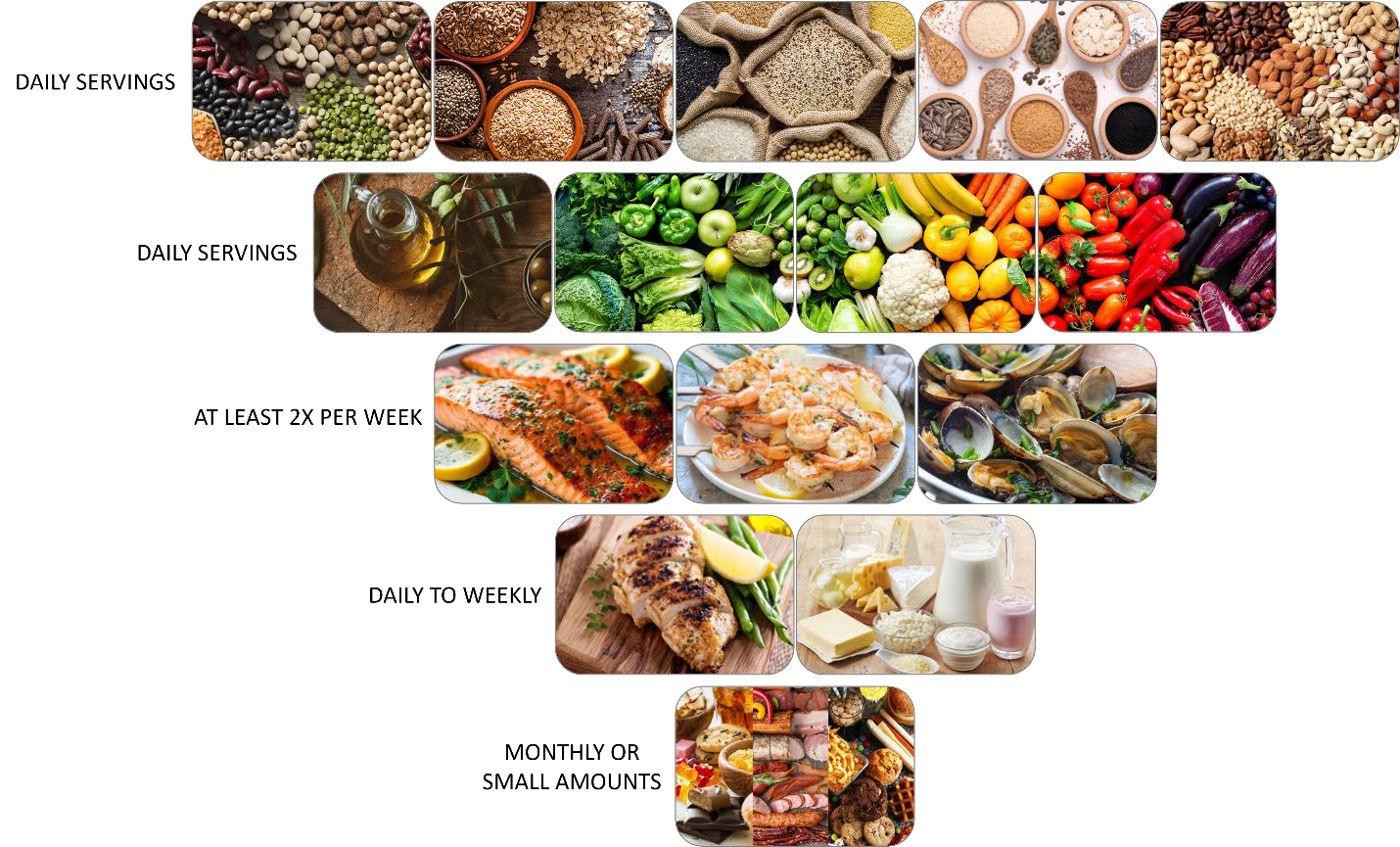

#### The Mediterranean Diet Overview

Base every meal around:

- Whole grains, beans, and nuts/seeds
- Vegetables and fruits (of different colors!)
- Olive oil as the primary source of fat Eat at least 2 times per week:
- Fish and seafood

Eat moderate portions daily to weekly:

- Chicken and turkey (not fried)
- Low-fat or nonfat dairy and cheeses
- Eggs

Eat less often (in very small amounts):

- Red meat (beef, pork, lamb) and processed meats (sausage, hotdog/corn dog, ham, bacon, and deli meat, jerky)
- Sweets, baked goods, desserts
- Ultra-processed foods (fast food, frozen meals, snacks, sugar-sweetened beverages)

#### Quick Changes to Make:

- Avoid processed meats (e.g. hot dog, deli meat, bacon, or sausage)
- Limit ultra-processed foods and sugar-sweetened beverages
- Have fruits and vegetables daily
- Add pulses (beans, lentils, peas) to your meals
- Choose fish and seafood, chicken, or turkey instead of red meat

#### Sample Meals:

BREAKFAST

- Oatmeal (not instant) with fresh berries and flaxseed or chia seeds
- Plain nonfat Greek yogurt with fresh berries and nuts
- Whole grain toast with hardboiled egg and ¼ avocado OR nut butter

LUNCH

- Baked salmon with kale quinoa salad* and roasted asparagus
- Bulgur black bean burger*on a whole wheat bun with cucumber and tomato salad*
- Apple tuna salad* with whole wheat pita bread or whole wheat crackers

DINNER

- Baked skinless, boneless chicken breast with baked rice and beans* and sautéed spinach (in olive oil)
- Salmon grain bowl*
- Black bean chili* with southern cornbread* and broccoli bean salad*
- Fish tacos* with cilantro lime crema* and roasted zucchini*
- Sweet potato and lentil soup*

SNACKS

- Unsalted/unseasoned nuts
- Fresh fruit

- Roasted chickpeas
- Whole grain crackers with classic* or edamame hummus*
- Raw veggies with white bean dip*

	Strawberry banana chia smoothie*

*Recipe available in the provided Recipe Book

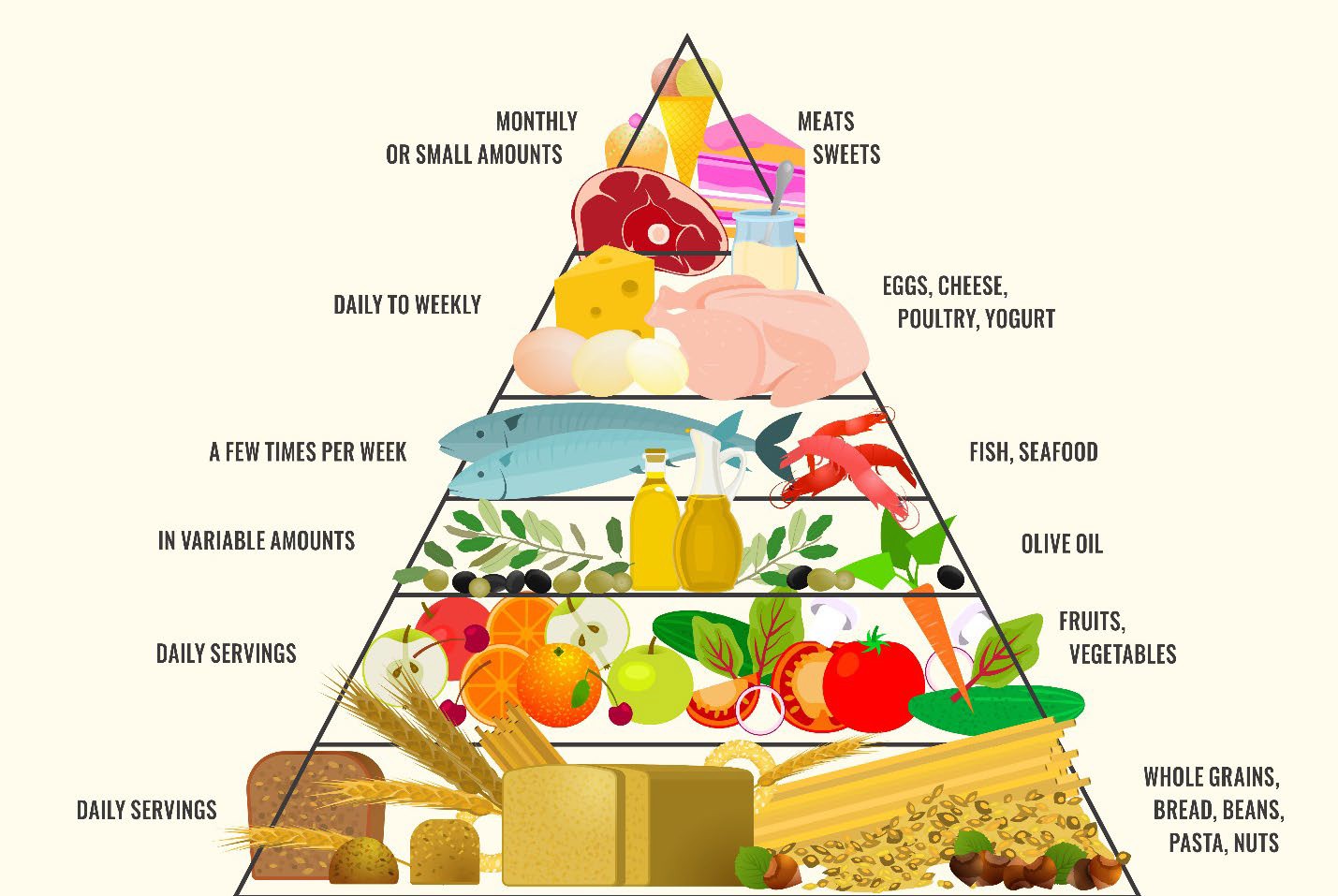

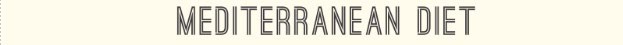

### Lesson 2: Fats

The Mediterranean diet is not a low-fat diet but emphasizes healthier types and sources of fat while limiting others. Fat is an important nutrient that your body needs for energy, protecting organs, and absorbing vitamins (A, D, E, K). Some fats are healthier than others. It is:

- Rich in omega-9 and omega-3 fatty acids, monounsaturated, and polyunsaturated fats found in plant foods and fish and seafood.
- Low in saturated and trans fats found in animal foods and processed or packaged foods.

Replacing unhealthy fats, like the fats from animal products, with healthier fats, like fats from plant sources, can lower your risk for major health problems.

#### Some Fats Are Healthier Than Others

| **FOOD SOURCES** | **SERVING SIZE** |
| --- | --- |
| **FATS TO EAT DAILY** | |
| Extra virgin olive oil* | 1 TBSP |
| Avocado / avocado oil | Avocado: ¼ medium Oil: 1 TBSP |
| **FATS TO EAT OFTEN** | |
| Seafood (sardines, anchovies, mackerel, herring, tuna, salmon, oysters) | 3 oz (~size of deck of cards) |
| Nuts (peanuts, almonds, hazelnuts, pecans, walnuts) and their nut butters | Nuts: ¼ cup  Nut butter: 2 TBSP |
| Seeds (pumpkin, sesame, sunflower) and their seed butters | Seeds: ¼ cup  Seed butter: 2 TBSP |
| Flaxseeds / Flaxseed oil | Flaxseed: 2 TBSP  Oil: 1 TBSP |
| Chia seeds | Chia seed: 2 TBSP |
| **FOODS WITH UNHEALTHY FATS TO AVOID** | |
| Red meat: beef, pork, and lamb | |
| Processed meat: sausage, hotdog/corn dog, ham, bacon, deli meat, jerky | |
| Poultry (including duck) with skin | |
| Dairy products: whole milk and products made with whole milk (cheese, heavy cream, half and half, butter, cream cheese, sour cream, ice cream) | |
| Coconut and coconut oil | |
| Palm oil and palm kernel oil | |
| Solid fats: stick margarine and vegetable shortening | |
| Commercially baked goods: chips, crackers, cakes, muffins, pie crusts, cookies, sweat breads/pastries | |
| Pre-mixed products: cake mix, pancake mix, Yoohoo chocolate drink, some hot cocoa mixes | |
| Packaged snack foods: crackers, microwave popcorn, chips, candy | |
| Fried foods: French fries, fried chicken, fried fish, chicken nuggets/strips, hard taco shells (including frozen store-bought types) | |

*Olive oil is a unique and important component of the Mediterranean diet. It is the primary oil used for cooking and “dressing” foods in place of butter and other oils.

#### Unhealthy fats include “saturated” and “trans fat.”

They are very common and many Americans eat them every day. However, these are not good for your heart, and may not be good in terms of prostate cancer. These fats are not part of the Mediterranean diet. We encourage limiting or avoiding these unhealthy fats as they are not part of the Mediterranean diet.

#### Goal

Aim for 4-5 servings of healthy fats daily. Avoid the “unhealthy” fats. Serving sizes for some foods that contain healthy fats are listed above.

#### Healthful Tips

The following tips can help you choose healthy fats over unhealthy fats which will help you limit the amount of dietary cholesterol you eat, too.

**STOP**

- Limit or avoid red meat (beef, pork, and lamb) and processed meats (sausage, hotdog/corn dog, bacon, deli meat, jerky).

**CHANGE**

- Bake with heart-healthy oils such as olive oil or avocado oil. You can also use 1 ½ cup ground flaxseed to replace ½ cup of butter, margarine, oil, or shortening (3:1 substitution ratio).
- Add a slice or smear of avocado to your sandwich instead of using mayonnaise or butter.
- Remove the skin from chicken and turkey before cooking.

**START**

- Always use olive oil when cooking, sauteing, pan-searing, or when making your own salad dressing. If cooking or roasting at high heat (>400°F), use avocado oil.
- Include nuts/seeds and nut/seed butters in snacks and meals.
  - Enjoy a 1-ounce serving of dry-roasted, unsalted nuts as a snack.
  - Natural nut butters without added salt, sugar, and palm oil are the best choices. Spread them on whole grain bread or fruits such as apples and bananas.
  - Add ground flaxseed or chia seeds along with berries to yogurt, oatmeal, or smoothies for a quick easy breakfast.
- Eat fish and seafood at least 2 times per week.

### Lesson 3: Fruits and Vegetables

Plant foods are the foundation of the Mediterranean diet. They fill you up with fewer calories. You should eat many fruits and vegetables each meal without changing or processing them (for example, don’t fry or juice). Increased intake of fruits and vegetables lowers risks of heart attacks and strokes and may prevent prostate cancer from worsening.

#### Vegetables

**GOAL**: Aim for 3-4 servings of vegetables daily.

| **FOOD SOURCES** | **SERVING SIZE** |
| --- | --- |
| **VEGETABLES TO EAT DAILY** | |
| Leafy greens: turnip, Bok choy, chard, collard, dandelion, kale,  mustard, spinach, endive, watercress, arugula, cilantro, dark green leafy mixed greens, romaine | 1 cup cooked OR  2 cups fresh |
| Broccoli and cauliflower, carrots, pumpkin, peppers, white and sweet potato, tomatoes, squash/zucchini, corn, peas, asparagus, cabbage, celery, cucumbers, green beans, mushrooms, okra,  onions, eggplant | 1 cup cooked |

**VEGETABLES TO AVOID**

100% vegetable juice – limit to 1 cup daily

Condiments “made” with vegetables (ketchup, relish, tartar sauce, wasabi)

Fried vegetables (any type such as French fries, hash browns, onion rings, fried okra, fried green tomatoes, fried Brussels sprouts, fried olives, fried cucumbers, fried yucca, potato/veggie chips/straws)

Vegetable dishes made with sauces, butter, gravies, added sugar, or with red/processed meats (such as scalloped potatoes/buttermilk mashed potatoes with butter/potato casseroles, corn pudding/corn casseroles/corn fritters, greens with bacon/ham hocks/smoked turkey, green bean casseroles, sweet potato casseroles/candied yams, creamed spinach/greens, potato salad)

#### Fruits

**GOAL**: Aim for 2.5 servings of fruits daily with at least 1 cup of berries each *week*.

| **FOOD SOURCES** | **SERVING SIZE** |
| --- | --- |
| **FRUITS TO EAT DAILY** | |
| Apple | 1 small, ½ large, 1 cup chopped or applesauce |
| Banana | 1 large, 1 cup sliced |
| Berries | 1 cup, fresh or frozen |
| Citrus fruits | 1 medium grapefruit, 1 large orange |
| Grapes, kiwi, mango, papaya, pineapple | 1 cup |
| Melons: honeydew, cantaloupe, watermelon | 1 cup diced or melon balls |
| Pear | 1 medium |
| Peach and Plum | 1 large peach, 2 large plums |

| Canned fruits in 100% juice (such as fruit cocktail, pears, peaches, mandarin oranges) | 1 cup |
| --- | --- |
| Dried fruits with added sugar (such as raisins, prunes, apricots, figs, dates, cranberries, blueberries, mangos) | ¼ cup |

**FRUITS TO AVOID**

100% fruit juice – limit to 1 cup daily

Canned fruits **in syrup** (such as fruit cocktail, pears, peaches, mandarin oranges)

Dried fruits **with added sugar** (such as raisins, prunes, apricots, figs, dates, cranberries, blueberries, mango)

Fried fruits (such as plantains)

Fruit-based foods made with sauces, butter, and added sugar (pies, jelly, jam, preserves, marmalade, compote with added sugar, candied fruit, fruit snacks, maraschino cherries, fruit- flavored yogurt/yogurt with real fruit)

#### Healthful Tips

Choose options that are full of nutrients and limited in added sugars, saturated fat, and sodium.

**STOP**

- Avoid canned or frozen fruit with added sugar or syrup (instead look for “in 100% juice” on the packaging).
- Limit fruit and vegetables juices to 1 cup daily. Whole fruits are better than drinking juice – contains more fiber which also helps you feel full.
- Avoid fried vegetables such as French fries, hash browns, onion rings, fried okra, fried green tomatoes, fried Brussels sprouts, fried olives, fried cucumbers, fried yucca, potato/veggie chips/straws.
- Limit adding sauces/dressings to vegetables such as salad dressings and queso (instead try hummus).
- Avoid dried fruit and fruit-based foods made with sauces, butter, and added sugar.
- Limit adding sauces/toppings to fruits such as chocolate syrup, sugar/honey/agave, and whipped cream.

**CHANGE**

- Spruce up your sandwich, wrap, or burritos by adding spinach, onions, lettuce, tomatoes, or shredded carrots.
- Make your salad colorful:
  - Add a variety of colored vegetables such as corn, radishes, red onions, tomatoes, red cabbage, bell peppers, etc.
  - Add grapefruit or tangerine sections, apples, or grapes in salads.
- Prepare vegetables without red meats and their fats; use olive oil and fresh herbs.
- Replace creamy dressings with olive oil-based vinaigrettes (for salads and coleslaw)
- Modify vegetable dishes made with sauces, butter, gravies, added sugar, or with red/processed meats to healthier versions such as sauté with olive oil or roasted in the oven.

**START**

- Start your day with fruits and or vegetables:
  - Add leftover cooked vegetables to an omelet or breakfast wrap or add spinach to smoothies.
  - Add your favorite seasonal or frozen fruit to (hot or cold) cereals or yogurt; top pancakes or waffles with bananas, berries, or chopped apples.
- Prepare fruits and vegetables ahead for easy snacking at home or on the go:
  - Baby carrots, celery sticks, jicama sticks, grape/cherry tomatoes, sugar snap peas, oranges, bananas, apples, and pears are all portable snacks when out.
  - Canned fruit cups packed in water are also great options.
- Try a vegetable stir-fry or roast vegetables using a mixture of your favorite vegetables.
- Try a dense bean salad to incorporate a variety of vegetables and fiber- and protein-packed beans.

### Lesson 4: Beans & Whole Grains

#### Beans, Peas and Lentils

Beans, peas and lentils (which go by the word "Pulses”) are an important source of plant-based protein in the Mediterranean diet – also providing excellent sources of dietary fiber, folate, potassium, iron, zinc, and protein. The high fiber and protein content in this food group keeps you full for longer. Pulses may decrease risk of cardiovascular disease and gut health.

#### Goal

Aim for 6 servings of pulses weekly. Each serving of pulses is **½ cup cooked**.

- Beans: black, black-eyed (cow) peas, fava, garbanzo, kidney, lima, mung, navy, pigeon, pink, pinto, white
- Peas: split green and yellow peas
- Lentils: red, brown, or green lentils
- Soy products: tofu, tempeh, edamame (without pods)

#### Whole Grains

In the Mediterranean diet, whole grains provide important nutrients while refined/processed grains are avoided. Whole grains do not undergo extensive processing, which removes nutrients like dietary fiber dietary fiber, iron, and B vitamins. Only foods made with 100% whole grains are considered whole grain foods. **Eating daily whole grains is a key part of the Mediterranean diet.**

#### Goal

Aim for 3 servings of whole grains daily. The amount of some common grains that count as 1 serving equivalents are below.

| **FOOD SOURCES** | **SERVING SIZE** |
| --- | --- |
| **WHOLE GRAINS TO EAT DAILY** | |
| 100% whole wheat breads (bread, pita bread, English muffin, bagel, buns, rolls) | 1 reg slice, ½ English muffin, mini 2” bagel, ½ reg bagel, ½ hamburger/hotdog bun or 6” roll |
| Corn and whole wheat tortillas | 1 small 6” tortilla |
| Ready-to-eat whole wheat cereals | 1 cup |
| Whole grains (oats, quinoa, millet, barley, buckwheat, bulgur, sorghum) | ½ cup, cooked |
| Brown and wild rice | ½ cup, cooked |
| Whole wheat pasta | ½ cup, cooked |
| Popcorn, plain | 3 cups, popped |
| Whole wheat crackers | 7 crackers |

**REFINED GRAINS TO AVOID**

White breads (sliced bread, pita, English muffin, bagel, buns, rolls)

White flour tortillas

Ready-to-eat cereals with lots of added sugar (Fruit Loops, Cinnamon Toast Crunch, Lucky Charms, Cap’n Crunch, Cocoa Puffs, etc.)

White rice

White/regular pasta

Regular crackers (Ritz, saltines, water crackers, Cheez-Its, Goldfish, graham crackers, pretzels,

etc.)

Other fried grain-based foods (such as tortilla/corn chips, hushpuppies, chimichangas, croquettes, etc.)

#### Healthful Tips

**STOP**

- Avoid ready-to-eat cereals with >5g of added sugar per serving. Choose whole grain ready-to-eat cereals without lots of added sugar.
  - Examples: Original Cheerios, Post Original Shredded Wheat, Uncle Sam Original Whole Wheat & Flaxseed Cereal, Fiber One Original Bran Cereal, Post Grape-Nuts Original, Total Whole Grain Cereal, Weetabix Whole Grain Cereal, Wheaties Whole Wheat Flakes
- Limit or avoid fried grain-based foods such as tortilla/corn chips, hushpuppies, chimichangas, croquettes, etc.

**CHANGE**

- Choose breads labeled as “whole wheat” instead of white breads (sliced bread, pita, English muffin, bagel, buns, rolls).
  - Multigrain is not the same as whole grain. Look for whole wheat flour as the first ingredient on the food label.
- Instead of white flour tortillas, switch to corn tortillas (or whole wheat flour tortillas) as a whole grain option.
- Choose whole grain crackers instead of regular crackers like Ritz, saltines, and Goldfish and Cheez-Its.

**START**

- Add whole grain foods like brown rice, quinoa, barley, farro, and whole wheat pasta to your diet.
  - Brown rice and whole wheat pastas may taste different than white rice and pasta; you can start by using half wheat/half white rice and pastas.
  - Try having whole grains like old-fashioned oats, buckwheat, farro, millet, quinoa for breakfast.
- Pulses are good sources of fiber and protein and can be a healthy, high fiber substitution for meat. They are also less expensive than meat!
  - Beans can be used in main dishes, side dishes, salads, pastas, dips and spreads, and baked goods. (see recipes in recipe book)
  - Soak dry beans overnight to shorten cooking time.
  - To reduce the sodium in canned beans, choose **low-sodium or no salt added**. Don’t forget to drain and rinse all canned beans.

### Lesson 5: Protein (Meats, Dairy, and Eggs)

Protein is a key part of the Mediterranean diet. Protein foods include poultry, red meats, dairy, eggs, seafood, pulses, nuts, and seeds. Seafood, pulses, nuts, and seeds are discussed in other sections of this handbook.

Protein foods (both animal and plant proteins) provide important nutrients such as B vitamins, vitamin E, iron, zinc, and magnesium and can help keep you full. In the Mediterranean diet, there are good protein sources and bad protein sources.

#### Protein Sources

Animal proteins are complete proteins, which contain all 9 essential amino acids.

- Poultry include chicken, turkey, duck, and quail.
- Red meats include beef, pork, and lamb.
- Processed meats include sausage, hotdog/corn dog, ham, bacon, and deli meat, jerky.
- Dairy foods include milk, yogurt, cheese, and other foods such as milk-based meal replacements.

#### Healthy proteins include plant-based proteins and lean sources of animal proteins.

| **FOOD SOURCES** | **SERVING SIZE** |
| --- | --- |
| **BEST/HEALTHIEST PROTEINS** | |
| Chicken (without skin, not fried) | 3 ounces, cooked (size of deck of cards) |
| Seafood (see Lesson 6) | 3 ounces, cooked |
| Pulses (see Lesson 4) | ½ cup, cooked |
| Nuts & Seeds (see Lesson 2) | Nuts/Seeds: ¼ cup Nut/Seed butter: 2 TBSP |
| **HEALTHY/GOOD PROTEINS** | |
| Low/Non-fat plain Greek yogurt | 1 cup |
| Eggs | 1 large egg, 2 egg whites, 4 tbsp liquid egg white |

| **FOOD SOURCES** | **SERVING SIZE** |
| --- | --- |
| **PROTEINS TO AVOID** | |
| Red meat: beef, pork, and lamb | 3 ounces, cooked |
| Processed meat: sausage, hotdog/corn dog, ham, bacon, deli meat, jerky | 3 ounces, cooked |
| Poultry with skin or fried | 3 ounces, cooked |
| Full-fat milk and yogurt | 1 cup milk and yogurt |
| Full-fat cheeses | 1.5 oz hard cheese, 1 oz processed (American) cheese, 1/3 cup shredded cheese, ½ cup ricotta cheese, 2 cups cottage cheese, 2 oz queso fresco |
| Other dairy products (heavy cream, half and half, butter, cream cheese, sour cream, creamers) | 1 TBSP |

Unhealthy proteins include proteins that contain high amounts of saturated fats and processed meats.

Evidence suggest increased red/processed meat and dairy intake may increase prostate cancer risk. Increasing data suggests plant-based diet may lower prostate cancer risk, thus adherence to the Mediterranean diet helps to limit intake of red/processed meat and full-fat dairy.

#### Goals

Get most of your protein from fish/seafood, pulses, some poultry, and nuts/seeds. Each serving of meat is 3 ounces.

- Limit poultry to 5 servings (15 oz total) or less each *week*.
- Limit eggs to no more than 3 servings each *week*.
- Limit red and processed meats to 3 servings (9 oz total) or less each *week*.
- Limit dairy to no more than 2 servings daily and no more than 4 servings of full-fat dairy products each

*week*.

- Limit butter and creams to no more than 5 servings (5 TBSP total) each *week*.

#### Healthful Tips

**STOP**

- Avoid red meats (beef, pork, and lamb) and processed meats (sausage, hotdog/corn dog, ham, bacon, deli meat, jerky). Limit 3 servings (9 oz total) *or less* per week.
  - Choose leaner cuts of red meats like sirloin, top round, or flank steak. Look on the label for lean meats that are at least 90% lean; >90% lean is even better. Choose beef cuts labeled ‘choice’ or ‘select,’ instead of ‘prime’ which usually has more fat.
  - Try having breakfast without meat some mornings.
  - Sliced fresh beef or pork are better than any type of lunch/deli meat.
- Limit dairy foods like heavy cream, half and half, butter, cream cheese, sour cream, creamers.
  - Greek yogurt is a good substitute for sour cream and cream cheese in recipes.
  - Olive oil is a good substitute for butter when cooking.
  - Use low-fat cream or skim milk as a substitute for heavy cream.

**CHANGE**

- Change the way you think about proteins.
  - Have smaller amounts when you eat red meat or only on special occasions.
  - Focus on plant-based protein sources like pulses, nuts, seeds, and whole grains.
- Choose chicken or turkey more often than red and processed meats.
  - Choose leaner poultry (chicken breast without skin).
  - Baked or broiled chicken will be healthier than fried chicken. Remove skin and chicken fat before cooking to decrease saturated fat.
  - Fresh chicken or turkey are better than chicken/turkey lunch/deli meats.
- Choose ***low-fat*** or ***fat-free*** forms for dairy foods instead of full-fat dairy foods.
  - Low-fat or fat-free milk instead of full fat milk, creamers, heavy cream, or half and half for coffee.
  - Low-fat or fat-free PLAIN Greek yogurt as a snack or for breakfast. Make sure to choose yogurt without flavor and added sugar (check nutrition label and ingredient list). Fermented dairies have other health benefits.
- Eggs can be a good source of protein.
  - Add chopped hardboiled eggs to salad or eat whole as a snack.
    - Make egg salad with hummus or mashed avocado as a healthier alternative to mayo in traditional egg salad.
  - Add vegetables to an egg scramble or egg “muffins”.

**START**

- Consider including two or more meatless meals in your weekly menu (Meatless Mondays): meatless chili with kidney or pinto beans; black bean enchiladas; beans and rice; veggie burgers; and chef salads with garbanzo or kidney beans.
  - Combine beans or nuts/seeds with whole grains (red beans and brown rice, hummus with whole wheat pita bread, or nut butter on whole wheat bread) for a meal with complete proteins.
  - Eating a variety of plant-based proteins throughout the day will provide you with adequate protein.
  - There are some plant proteins that are complete protein sources, such as whole sources of soy (tofu, edamame, tempeh, miso, soymilk), buckwheat, and quinoa.
- Vary your protein choices by including (non-fried) seafood more often: fish (salmon, tuna, herring, trout, and tilapia) and shellfish (shrimp, crab, and oysters) are great choices.
- Add nuts for additional protein and reduce meats in a meal. (examples: slivered almonds on steamed vegetables, toasted peanuts or cashews in a vegetable stir-fry, walnuts or pecans to salads).
- If you don’t consume dairy products, look for calcium-fortified foods.

### Lesson 6: Fish and Seafood

Fish and seafood are important components of the Mediterranean diet. They are great sources of protein in the diet and help to keep you full. They are also good sources of other nutrients like iron, iodine, choline, vitamin B12, vitamin D, and selenium. Research has shown that substituting red and processed meats with fish and seafood is beneficial for health. Please keep in mind some types of fish are better than others.

#### Types of Seafood

*Each serving of seafood is 3 ounces (about the size of a deck of cards).

| **TYPE** | **SEAFOOD** |
| --- | --- |
| **FISH AND SEAFOOD TO EAT OFTEN** | |
| Fatty Fish | Sardines, anchovies, mackerel, herring, salmon |
| Lean Fish | Tilapia, trout, bass, cod, haddock, hake, pollock, “light”/yellowfin and “white”/albacore tuna |
| Crustaceans | Shrimp, crawfish, crab, lobster |
| Mollusks | Scallops, clams, oysters, mussels |

**FISH AND SEAFOOD TO AVOID**

Fried fish and seafood (fried fish, fried oysters, fried shrimp/crawfish, fried crab, fried clams, calamari)

Fish high in mercury (swordfish, marlin, shark, tilefish, King mackerel, bluefin/bigeye tuna)

#### Goal

Aim for at least 2 servings (6 oz total) of seafood each week. More is better.

#### Healthful Tips

**STOP**

- Avoid or limit your intake of fried fish and seafood and or seafood dishes with creamy sauces.

**CHANGE**

- Substitute red and processed meat in some recipes with fish or seafood.
  - Marinate and bake fish instead of grilling burgers.
  - Pan-fry white fish instead of making a steak.
  - Roast or grill salmon instead of beef, lamb, or ham.
  - Add grilled fish or shrimp instead of grilled chicken or steak to a salad.
  - Use canned tuna or salmon for sandwiches or grain bowls in place of deli meat.

**START**

- If you are not used to eating fish or seafood, start with milder tasting fish such as white fish (tilapia, cod, pollock). Adding acid (lemon and vinegar/wine-based marinades and sauces) can help bring out a bright, clean flavor.
- Try baked salmon, shrimp stir-fry, fish tacos, or clams with whole wheat pasta.
- Cook extra of your favorite fish and use the leftovers for another meal or two – a great way to get your seafood several times a week!
- Keep frozen fish in the freezer as a cost-effective option. You can cook from frozen or thaw fillets in about 15-20 minutes. Buy in bulk and freeze.
- Canned salmon, tuna, sardines, anchovies are easy and quick to use in recipes. Look for low-sodium options.

### Lesson 7: Eating Out

Eating out is a way of life for many people as it is easy and convenient to order out or get fast foods. Keep in mind though that many foods at restaurants and food trucks are not part of the Mediterranean diet. Foods prepared outside of the home are often high in calories, sodium, unhealthy fats, and don’t align with the Mediterranean diet. A diet high in these types of foods can place you at an increased risk of a heart attack or stroke. Keep in mind The New American Plate (shown below). Choose 2/3 or more vegetables, fruits, whole grains, and beans. Choose 1/3 or less animal protein.

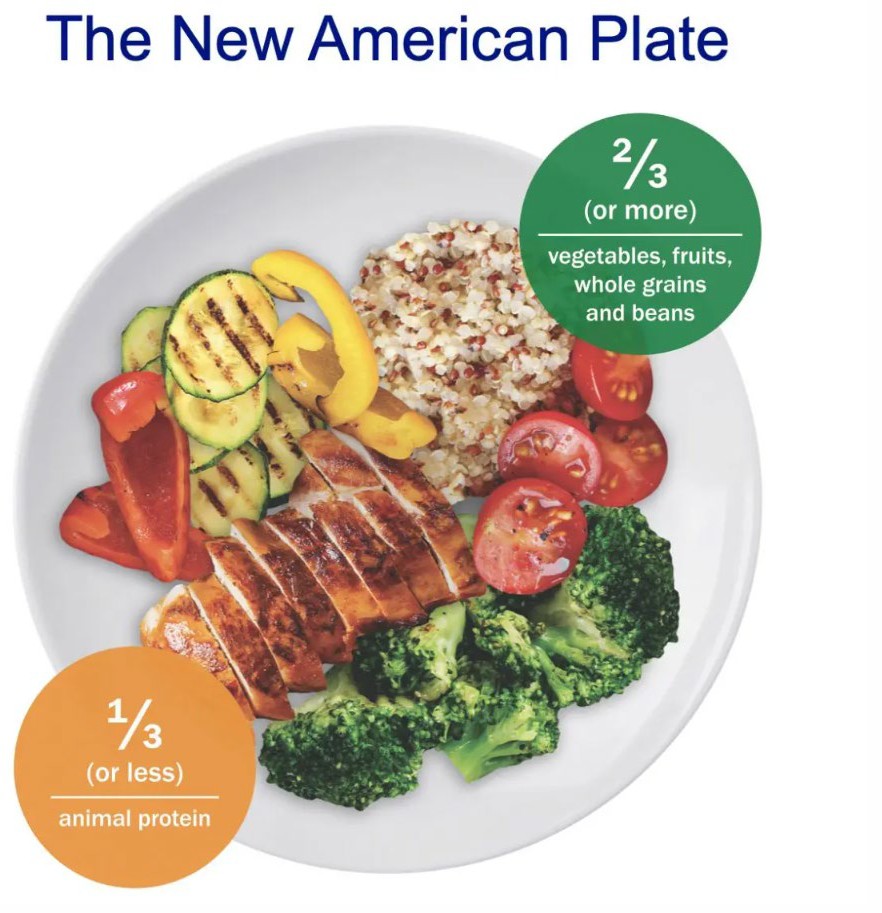

#### Healthful Tips

Having a plan will help you prepare for difficult situations and make healthy choices.

- Before eating at the restaurant:
  - Select restaurants that offer low-fat, low-calorie choices and meals with lean protein sources on the menu. You can check their websites to find out.
  - Avoid going to a restaurant when you’re starving.
- When at the restaurant:
  - Be the first to order. You will be less likely to order unhealthy meals that other people order.
  - Keep food off the table that you do not want to eat. Ask the server to remove bread and butter from the table.
  - Look for dishes that highlight vegetables. Start your meal with the lowest calorie items first such as a salad or vegetables.
  - Order salad dressing, gravy, sauces, or spreads on the side.
  - Split a main dish with someone or before starting your main dish, put the amount you do not want to eat at that meal in a container to take home.
  - If having dessert, split it with someone or select fruit.
  - Choose water or unsweetened tea instead of soft drinks.
  - Eat slowly. Put your fork down between bites or take a sip of water.
  - Ask yourself “Am I still hungry?” when you have eaten half of your meal. Often the answer is “no.” Save the food for your next meal.
  - Chew on gum or have a mint after you’re done eating.
- Keep a list of healthy dishes and restaurants. Refer to the list when eating out next.
- For parties or dinner parties, bring a healthy, low- fat and low-calorie dish to share with others.

#### Choose Your Foods Carefully

Look for these keywords on the menus when eating out.

| **LOWER FAT OPTIONS TO CHOOSE:** | **HIGH FAT OPTIONS TO AVOID/LIMIT:** |
| --- | --- |
| - Baked - Broiled - Boiled - Grilled - Poached - Roasted - Sauteed - Stir-fried | - Au gratin - Breaded - Buttery or buttered - Cheese sauce - Creamed, creamy, cream sauce - Fried, deep fried, batter fried, pan- fried - Gravy - Hollandaise - Scalloped, escalloped - Southern style |

Review these tips to choose healthier options at each restaurant type.

| **CHOOSE THESE:** | **AVOID/LIMIT THESE:** |
| --- | --- |
| **Pizza** | |
| - Half the cheese or low-fat cheese - Vegetable toppings such as onions, green peppers, mushrooms, tomatoes, etc. - Thin crust pizza | - Meat toppings such as pepperoni, bacon, or sausage - Extra cheese - Stuffed crust pizzas - Bread sticks, hot wings, and desserts |
| **Fast Food Chain** | |
| - Grilled, broiled, or roasted chicken without sauce - Turkey burger without cheese - Small side salad with vinaigrette dressing on the side | - Large hamburgers and cheeseburgers - Bacon - French fries - Fried chicken or fried fish - Sauces made with mayonnaise or cheese |
| **Sandwich** | |
| - 6” sub, whole wheat bread - Broth-based soups such as chicken noodle soup or vegetable soup - Fresh grilled chicken or turkey - Low-fat cheese | - Footlong sub - Creamy soups (and with potato, bacon, or cheese in the name) - Ham, deli turkey or chicken, tuna salad, bacon, meatballs, or steak |

| - Extra vegetable toppings - Mustard | - American cheese - Mayo or other sauces |
| --- | --- |
| **Mexican/Tex Mex** | |
| - Corn tortillas (not fried) - Grilled chicken fajitas - Salsa and guacamole - Grilled or roasted vegetables | - Enchiladas, chimichangas - Queso, chili con queso - Fried tortillas (crispy tacos), tortilla chips - Sour cream |
| **Chinese & Japanese** | |
| - Stir-fried chicken and vegetables - Steamed brown rice - Broth-based soup - Non-fried fish and seafood | - Teriyaki sauce - Fried rice or noodles - Egg rolls - Fried wonton - Tempura |
| **Italian (see pizza tips above)** | |
| - Whole wheat pastas - Pesto - Minestrone soups - Salads with dressing on the side - Seafood | - Sausage - Lasagna, manicotti, scampi, other pastas with a lot of cheese, cream, or butter - Fried or breaded meats or vegetables such as chicken parmesan and mozzarella sticks |
| **Seafood** | |
| - Broiled, baked, or boiled seafood with limited butter or sauces - Sides with vegetables and whole grains | - Fried seafood (including calamari) - Sides like hush puppies, French fries |
| **Steakhouse** | |
| - Shrimp cocktail - Broiled or grilled chicken and fish - Sides with vegetables and whole grains | - High fat cuts of steak - Mashed potatoes, potatoes au gratin - Sides like onion rings, French fries, and other fried vegetables |

#### Limit Eating Out

Bring your own lunch and snacks to work or on-the-go to avoid having to eat out during the day. Prepare meals ahead of time that can be heated up easily or packed with ice packs if a refrigerator and microwaves are not available. Prepare healthy snacks ahead of time (see Lesson 8) so that you can grab and go and resist reaching for a bag of commercially packaged snacks.

#### Sodium

Sodium is another word for salt. Most Americans’ diet includes too much sodium. Excess sodium intake can lead to high blood pressure, heart disease, and stroke. Most of the sodium in the diet comes from commercially processed/prepared foods and some sodium comes from salt used in cooking or at the table. Aim to have no more than 2,300 mg sodium daily.

#### Healthful Tips

While it can be hard to track salt intake throughout the day, here are tips to decrease sodium in your diet.

- Cut back on prepared foods that are high in sodium.
  - Read the Nutrition Facts label on packaged foods. Choose foods with 140 mg of sodium or less per serving.
  - Even processed foods that don’t taste salty (like breakfast cereal, bakery pastries, energy/sports drinks) are high in sodium.
- Eat smaller portions of salty foods.
  - Top sources of sodium in the American diet per the CDC: breads and rolls, lunch/deli meats, pizza, processed meats, soups, sandwiches, cheese, packaged savory snacks (crackers, chips, etc.).
  - Foods that are pickled, brined, or cured are usually high in sodium.
  - Commercially produced sauces, gravies, salsas, condiments, soy sauce, marinades, dressings tend to be high in sodium. You can make these at home to control how much salt is added.
- Look for canned beans and vegetables labeled “no salt added”. Make sure to rinse and drain all canned beans and vegetables.
- Flavor foods with fresh or dried herbs and spices instead of salt. Watch for store-bought herb/spice blend- they often contain salt. Make your own at home without the salt!

### Lesson 8: Snacks, Desserts, and Beverages

#### Why and How We Snack

Snacks can be a part of the Mediterranean diet if the right choices are made. Snacks were invented by the food industry in the mid-20^th^ century with the goal to sell cheap, tasty food that Americans will buy again and again. Well, that worked! A snack is generally defined as any food eaten between main meals. Most commonly, snacking happens when we become hungry between meals, need an energy boost, or simply because we like the taste of certain snack foods.

#### Can I include snacks, desserts, and beverages in a healthy diet?

Typical snacking patterns are not consistent with the Mediterranean diet. Snacks have been associated with both weight gain and weight maintenance, as well as with lower and higher diet quality. Although snacks can lead to health problems, snacks can be a regular part of a healthy diet. It depends on the following snacking behavior:

- What you snack on?
- Why you snack?
- How often you snack?
- How snacks fit into your overall eating plan?

Some benefits of snacking include providing a boost of energy if blood glucose levels drop between meals, helps curb appetite to prevent overeating at the next meal, provides extra nutrients when choosing certain snacks, and can help to maintain adequate nutrition if having poor appetite and cannot eat full meals (such as due to an illness).

#### How to include snacks, desserts, and beverages in a healthy diet

Studies show that snacking on whole foods containing protein, fiber, and whole grains enhance satisfaction. A general rule of thumb is to aim for 150-200 calories per snack – should be enough to satisfy but not interfere with your appetite for the next meal.

**FOOD**

**BEST/HEALTHIEST SNACKS, DESSERTS, BEVERAGES**

Beverages: water, coffee/tea (without added sweeteners and creamers)

Fruits and vegetables without added sugar, toppings, and sauces/dressings

Nuts/seeds and nut/seed butters

Plain popcorn

High protein, low fat dairy (i.e. nonfat plain Greek yogurt)

**SNACKS, DESSERTS, BEVERAGES TO AVOID**

Beverages high in added sugars: regular sodas, sports beverages, energy drinks

Alcoholic beverages

Commercially made pastries/desserts: doughnuts, churros, sweet rolls, pies, cakes, bakery cookies, candy bars, cream-filled desserts, ice cream (bars)

Commercially made snacks: chips, crackers (i.e. Ritz), cookies (i.e. Oreos, Chips Ahoy), granola bars (i.e. Nutri-grain Bars, Quaker Chewy Dipps Bars, Fiber One Chewy Bars, Special K Chewy Bars, some Larabars, some KIND bars, some Nature Valley bars, some Kashi bars)

#### Alcohol

Research shows that drinking any amount of alcohol increases risk for cancers. To be consistent with the Mediterranean diet, have no more than 1 drink per day for women and up to 2 drinks per day for men of wine (not beer or liquor). A drink is defined as 12 ounces of beer, 8-9 ounces of malt liquor, 5 ounces of wine, and

1.5 ounces of hard liquor (see picture). If you do not drink, do not start.

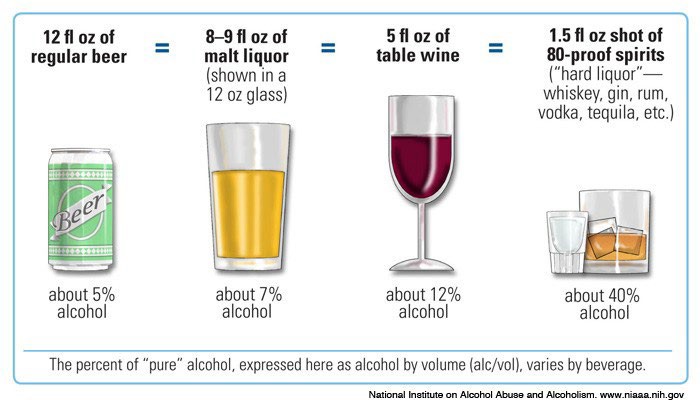

#### Healthful Tips

Snacks, desserts, and beverages are often high in added sugar, sodium, saturated fat, and/or non-nutritious calories. Try these tips below for healthier snacking.

**STOP**

- Limit commercially made pastries/desserts such as doughnuts, churros, sweet rolls, pies, cakes, bakery cookies, candy bars, cream-filled desserts, ice cream (bars) as they are high in added sugar, sodium, and saturated fats (and calories).
  - Save sweets and desserts for a special treat or celebration.
- Commercially made snacks that contain high fat, sodium, and added sugars.
- Avoid sugar-sweetened beverages and limit beverages containing low-calorie sweeteners; sweeteners include high fructose corn syrup, other syrups, sucrose, malt, fructose, and sugar alcohols.
- It is best not to drink alcohol. If you do drink, have no more than 2 drinks (12oz beer, 5oz wine, or 1oz liquor) for men and no more than 1 drink for women.
  - Be aware that restaurants and bars often serve larger than standard size alcoholic drinks.
  - Sip slowly and make one drink last a long time.
  - Consider alternating alcoholic and nonalcoholic drinks.

**CHANGE**

- Substitute sugar-sweetened beverages and low-calorie-sweetened beverages with options below:
  - Water or carbonated, unsweetened, flavored water
  - Unsweetened tea or plain coffee
  - Low-fat or fat-free milk
  - Add flavor to water with lemon, lime, or orange slices; can also try mint leaves or fresh/frozen berries if plain water is not palatable.
- Try fruit for dessert such as a fresh fruit salad, baked apples/peaches with cinnamon, or just a piece of fruit.
- Share a dessert with a friend/family. Eat only half a serving for half of the added sugars, unhealthy fats, and calories for each of you.

o

**START**

- Use “mindfulness” strategies to help reduce over-snacking:
  - Think ahead of time about snack choices.
  - Pre-portion out snacks into a small bowl or container.
  - Take the time to enjoy small bites
  - chew thoroughly, using the senses to fully appreciate the textures and tastes of the snacks.
- Choose snacks that will be satisfying. Consider these snack choices depending on your craving/preference:
  - Crunchy: raw vegetables sticks, nuts, seeds, whole grain crackers, apple, popcorn
  - Creamy: low-fat cottage cheese, Greek yogurt, hummus, avocado
  - Sweet: fresh fruit, dark chocolate (>70% cocoa)
  - Savory/Salty: roasted chickpeas, nuts, nut butters
- Build satisfying snacks with different food groups!
  - Greek yogurt + berries
  - Apple or banana + nut butter
  - Whole grain cracker + avocado
  - Vegetable sticks + hummus or guacamole
  - Unsalted nuts/seeds + raisins or other dried fruits + plain popcorn
  - Fruit + nuts + dark chocolate
- Coffee
  - It’s ok to drink coffee!
  - Try to avoid adding milk, cream or sugar

### Sources

The Nutrition Source, Harvard T. H. Chan School of Public Health MyPlate, USDA

Nutrition Care Manual (Academy of Nutrition and Dietetics) AICR
